# Moral injury and psychological wellbeing in UK healthcare staff

**DOI:** 10.1101/2022.06.16.22276476

**Authors:** Victoria Williamson, Danielle Lamb, Matthew Hotopf, Rosalind Raine, Sharon Stevelink, Simon Wessely, Mary Docherty, Ira Madan, Dominic Murphy, Neil Greenberg

## Abstract

**Background:** Potentially morally injurious events (PMIEs) can negatively impact mental health. The COVID-19 pandemic may have placed healthcare staff at risk of moral injury.

**Aim:** To examine the impact of PMIE on healthcare staff wellbeing.

**Method:** 12,965 healthcare staff (clinical and non-clinical) were recruited from 18 NHS-England trusts into a survey of PMIE exposure and wellbeing.

**Results:** PMIEs were significantly associated with adverse mental health symptoms across healthcare staff. Specific work factors were significantly associated with experiences of moral injury, including being redeployed, lack of PPE, and having a colleague die of COVID-19. Nurses who reported symptoms of mental disorders were more likely to report all forms of PMIEs than those without symptoms (AOR 2.7; 95% CI 2.2, 3.3). Doctors who reported symptoms were only more likely to report betrayal events, such as breach of trust by colleagues (AOR 2.7, 95% CI 1.5, 4.9).

**Conclusions:** A considerable proportion of NHS healthcare staff in both clinical and non-clinical roles report exposure to PMIEs during the COVID-19 pandemic. Prospective research is needed to identify the direction of causation between moral injury and mental disorder as well as continuing to monitor the longer term outcomes of exposure to PMIEs.

## Introduction

Many professionals face ethical decision-making dilemmas during their careers. It is increasingly recognised that making ethical judgements in complex circumstances, events where someone’s moral or ethical code has been broken, can contribute towards the development of *moral injury* [1,2]. Moral injury is the psychological distress experienced following an event which violate one’s moral beliefs or expectations [2]. Morally injurious events typically include acts of commission (e.g. deliberate negligence or mistreatment of others), omission (e.g. witnessing unethical behaviour of others and failing to intervene), and betrayal by trusted others (e.g. was not provided with adequate support). Experiences of moral injury can lead to maladaptive beliefs that threaten one’s perceived identity or view of the world – for example, ‘I’m a terrible person,’ ‘I failed my team,’ ‘my colleagues don’t care about me’ – as well as feelings of guilt, shame, disgust, and anger [3]. While moral injury is not a mental health diagnosis, these post-event changes in appraisals and affect are associated with the development of mental health disorders. Extensive studies have found significant associations between reports of exposure to potentially morally injurious events (PMIEs) and mental health disorders, including posttraumatic stress disorder (PTSD), depression, and suicidality [4,5]. Identified risk factors for heightened distress following PMIEs include feeling psychologically or emotionally unprepared for the event; concurrent exposure to other stressors (e.g. serious illness); a perceived lack of support from those in a more senior role; or PMIEs involving vulnerable victims (e.g. children, elderly) [3]. Whereas factors such as leaders taking responsibility for events and empathetic support from colleagues have been found to be protective [3]. Whilst much of the research carried out to date has examined experiences of moral injury in military contexts (e.g. [5,7,8]); the COVID-19 Pandemic has raised awareness that healthcare staff may also be at riskof moral injury and the pandemic may have increased the risk of healthcare staff exposure to PMIE’s [9]. Healthcare staff may, not unreasonably, expect their employer to provide them with adequate protection/resources to fulfil their roles safely. Moral injury may therefore follow situations in which healthcare workers cannot provide good quality care because of lack of resources, or where staff perceive they received inadequate training or adequate personal protective equipment (PPE) [10,11].

Studies in other occupational groups such as military personnel, have identified contextual risk factors– including experiences of poor leadership, low morale and unit cohesion– are associated with an increased likelihood of moral injury and mental ill health [13–15]. Whether such factors are also applicable to healthcare staff is less well understood. Moreover, longitudinal studies carried out during the 2020-2021 COVID-19 Pandemic, indicate that healthcare workers may be vulnerable to particular types of distress. For example, National Health Service (NHS) Intensive Care Unit (ICU) nursing staff have been found to be particularly likely to experience mental health difficulties, including suicidal ideation, compared to their doctor or non-clinical counterparts [16]. This may be due to factors related to their role (e.g. nursing staff spend more time with patients but have less control over their care) or demographics (e.g. typically less well paid) [17]). It is also possible that healthcare staff may be exposed to different types of PMIEs (e.g. commission, betrayal, witnessing), with some events experienced as more distressing than others. Nonetheless, whether healthcare staff are more likely to experience certain types of PMIEs and suffer with specific moral injury-related mental health difficulties is unclear.

The aim of this study was to examine the experience and impact of PMIEs and moral injury-related mental health difficulties in a nationally representative sample of NHS-healthcare staff. Our study draws on NHS CHECK data [18], which includes clinical as well as non-healthcare staff, and measures moral injury as well as common mental disorders (anxiety, depression), alcohol misuse and PTSD symptoms. Here we report the prevalence and nature of PMIEs in nursing, medical and non-clinical staff as well as the association between PMIE exposure and adverse mental health symptoms.

## Methods

### Ethical approval

Ethical approval for the study was granted by the Health Research Authority (reference: 20/HRA/210, IRAS: 282686) and local Trust Research and Development approval. The study was approved as having Urgent Public Health Status by the NIHR in August 2020. The study was NIHR CRN adopted (ID CPMS ID 46176).

### Participants

Participants were drawn from the NHS CHECK cohort, which consists of clinical and non-clinical staff from 18 NHS Trusts in England. Trusts were purposively selected for inclusion in the study in order to provide a range of sizes, locations, and types (mental health and acute). All staff in participating Trusts were eligible for inclusion in the study, including full- and part-time staff, clinical and non-clinical, fixed-term, permanent, and bank staff. The cohort is described in more detail elsewhere [18,19].

### Recruitment

All staff in participating Trusts were invited to participate in the study via emails sent by senior Trust managers, chief nursing officers, medical directors, occupational health departments, and trade union representatives. The study was discussed in team meetings and briefings, advertised in newsletters and on Trust intranets, and via screen savers on Trust computers. Prize draw incentives were offered (10x£50 gift vouchers and 10x£250 gift vouchers).

### Procedure

Data were collected via online surveys, developed using Qualtrics software. The survey included an information sheet (confirming participation was voluntary and confidential), and consent form, and the subsequent survey questions took around 5-10 minutes to complete. At the end of the survey, participants were offered the opportunity to complete an additional, longer, survey, which took around 10-15 minutes to complete.

### Materials

The surveys included the following validated measures:

### Moral Injury Events Scale (MIES)

A 9-item scale asking whether respondents experienced events that conflicted with their own moral values [20]. Responses are on a Likert scale of 1 (strongly disagree) to 6 (strongly agree), with a total score range of 9-54, and a higher score indicating greater exposure to potentially morally injurious events (PMIEs) (see Table 4). We dichotomised the scale by using moderately agree or strongly agree on one or more item to indicate exposure to PIMEs [21].

### Post-Traumatic Stress Disorder checklist (civilian version) (PCL-6)

A 6-item scale consisting of questions about PTSD symptoms [22]. A Likert scale of 1 (not at all) to 5 (extremely) is used, with a total score range of 6-30, higher scores indicating more symptoms of PTSD, and a cut-off score of 14 or more indicating the presence of probable PTSD.

### General Health Questionnaire (GHQ-12)

The 12-item version of this scale was used, which asks about general psychological distress [23]. A Likert scale of 0 (better than usual) to 3 (much worse than usual) is used, and the GHQ scoring method is used, where 0 and 1 are scored as 0, and 2 and 3 are scored as 1, giving a range of 0-12 for total score, where higher scores indicate more distress. A cut-off score of 4 or more indicates presence of probable common mental disorder (CMD).

### Generalised Anxiety Disorder (GAD-7)

A 7-item scale consisting of questions about symptoms of anxiety [24]. A Likert scale of 0 (not at all) to 3 (nearly every day) is used to give a total score range of 0-21, with higher scores indicating more symptoms of anxiety disorder. A cut-off score of 10 or more indicates presence of probable anxiety disorder.

### Patient Health Questionnaire (PHQ-9)

A 9-item scale which asks questions about symptoms of depression [25]. A Likert scale of 0 (not at all) to 3 (nearly every day) is used to give a total score range of 0-27, with higher scores indicating more symptoms of depression. A cut-off score of 10 or more indicates presence of probable depression.

### Alcohol Use Disorder Identification Test (AUDIT)

A 10-item scale that measures alcohol consumption [26]. There is an initial screening question of whether respondents ever drink alcohol, and then 10 questions assessing how often respondents engage in hazardous drinking behaviours, with a mixture of binary and Likert scale (0 – never, to 4 – daily/almost daily) responses available, resulting in a total score range of 0-40. Higher scores indicate more hazardous drinking behaviours, and a cut-off score of 8 or more indicates hazardous drinking.

### BAT-12

A 12-item scale consisting of questions about symptoms of burnout [27]. A Likert scale of 1 (never) to 5 (always) is used to give a total score, which is then divided by 12, giving a total score range of 1-5, with higher scores indicating more symptoms of burnout. A cut-off score of 3.02 indicates probable burnout.

### Data analysis

The measure of moral injury used in this study (the MIES) was part of the longer survey, and therefore only participants who completed both the short and long surveys are included in the dataset used here (N=12,965). Differences between those completing the long and short surveys are given in detail elsewhere [19]. Briefly, women, White participants, those in a relationship, those who were nurses or in non-clinical roles, and those who had not been redeployed, were more likely to complete the long survey.

The data used in these analyses were weighted, using Trust population demographic data obtained from Trust HR departments, in order to obtain estimates that better represent the population our sample is drawn from. Response weights were generated using a raking algorithm based on age, sex, ethnicity, and role, with missing data imputed using multiple imputation. Imputed data was used only for the purposes of weighting the data, as missing data was no more than 2% in any of the weighting variables [28].

We analysed the data in several stages. Firstly, we described the demographics of the cohort using frequencies and weighted percentages for categorical variables and means and standard deviations for continuous variables. Secondly, we described the demographics of those meeting and not meeting the cut-off for a binary construction of the MIES (those answering ‘moderately/strongly agree’ on one or more item, as used by [21]). Thirdly, we explored associations between MIES total score and the mental health symptom measures, and between the MIES subscales and the mental health symptom measures, using multilevel logistic regression models (adjusting for factors with statistically significant associations with MIES cut-off: age, sex, ethnicity, role, change in role, contact with COVID-19 patients, adequate PPE, adequate support from colleagues, managers, and family/friends, colleague died from COVID-19), with Trust as the grouping variable. Finally, we explored associations, by role, between those meeting cut-off on one or more of the mental health measures, and the MIES items, using multilevel logistic regression models (adjusting for the same factors as above) and using Trust as the grouping variable [29].

All analyses were carried out in Stata software [30].

## Results

Overall, of the 12,965 participants, 851 (9%) were doctors, 3,456 (31%) were nurses, 3,730 (32%) were other clinicians, while 4,693 (28%) were non-clinical staff (Table 1). The majority of participants were female (77%) and White (82%), with a mean age of 43 years (IQR 34-54). In terms of moral injury, 28% of the sample endorsed (moderately or strongly agreed with) one or more items of the MIES, which we refer to as meeting cut off (see Table 2). Acts of betrayal were most frequently reported across the sample (22% met cut off on the betrayal subscale), followed by acts of omission (15% met cut off on the omission subscale) and commission (6% met cut off on the commission subscale).

**Table 1.**
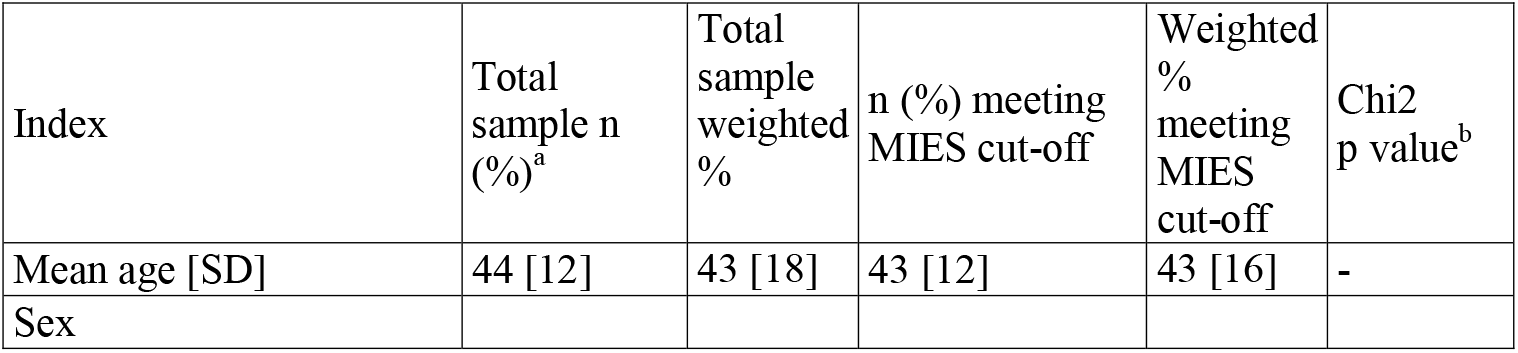
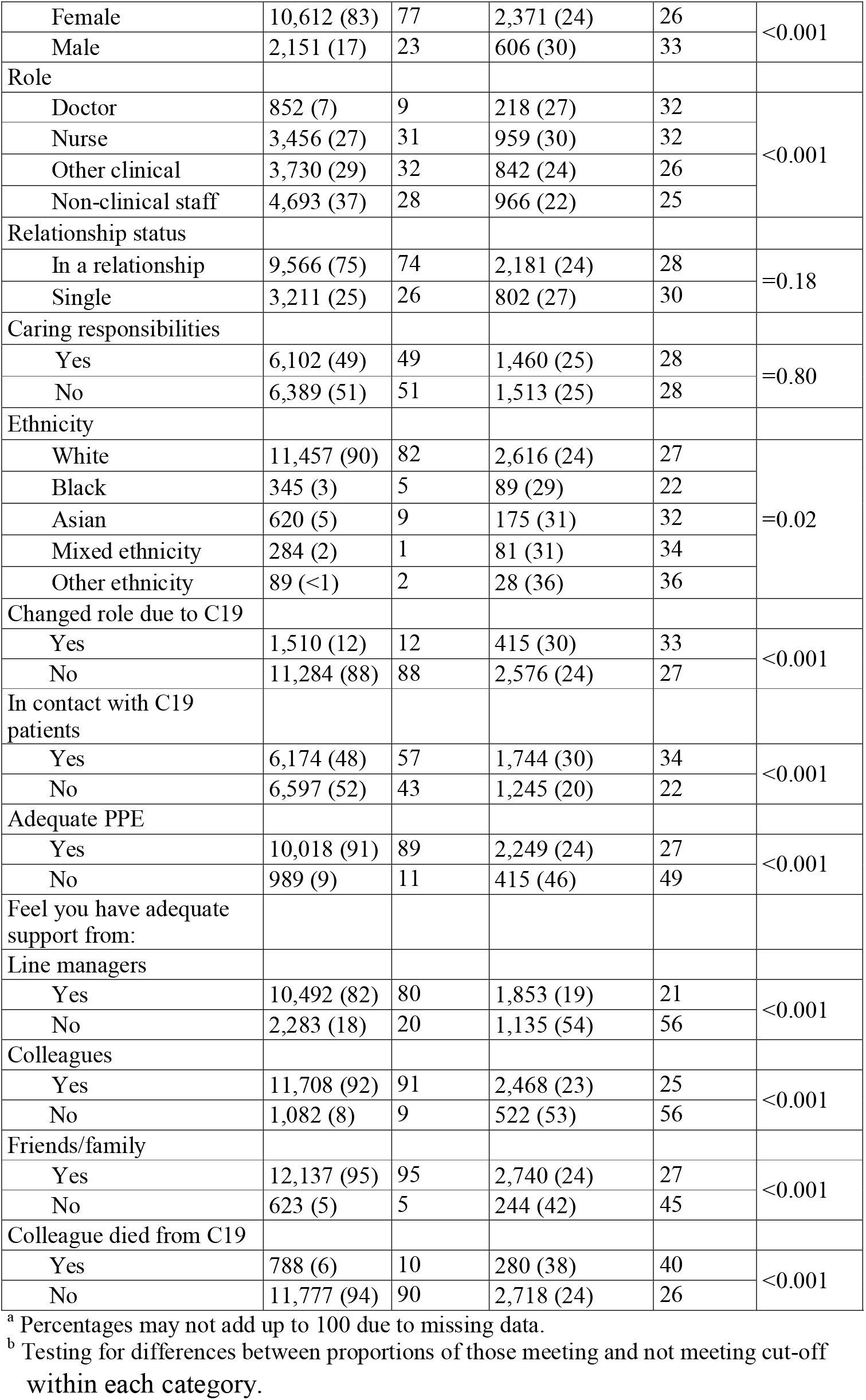
Participant demographic characteristics, and proportions of each demographic meeting MIES cut-off

| Index | Total sample n (%)<sup>a</sup> | Total sample weighted % | n (%) meeting MIES cut-off | Weighted % meeting MIES cut-off | Chi2 p value<sup>b</sup> |
| --- | --- | --- | --- | --- | --- |
| Mean age [SD] | 44 [12] | 43 [18] | 43 [12] | 43 [16] | - |
| Sex |  |  |  |  |  |
| Female | 10,612 (83) | 77 | 2,371 (24) | 26 | <0.001 |
| Male | 2,151 (17) | 23 | 606 (30) | 33 |  |
| Role |  |  |  |  |  |
| Doctor | 852 (7) | 9 | 218 (27) | 32 | <0.001 |
| Nurse | 3,456 (27) | 31 | 959 (30) | 32 |  |
| Other clinical | 3,730 (29) | 32 | 842 (24) | 26 |  |
| Non-clinical staff | 4,693 (37) | 28 | 966 (22) | 25 |  |
| Relationship status |  |  |  |  |  |
| In a relationship | 9,566 (75) | 74 | 2,181 (24) | 28 | =0.18 |
| Single | 3,211 (25) | 26 | 802 (27) | 30 |  |
| Caring responsibilities |  |  |  |  |  |
| Yes | 6,102 (49) | 49 | 1,460 (25) | 28 | =0.80 |
| No | 6,389 (51) | 51 | 1,513 (25) | 28 |  |
| Ethnicity |  |  |  |  |  |
| White | 11,457 (90) | 82 | 2,616 (24) | 27 | =0.02 |
| Black | 345 (3) | 5 | 89 (29) | 22 |  |
| Asian | 620 (5) | 9 | 175 (31) | 32 |  |
| Mixed ethnicity | 284 (2) | 1 | 81 (31) | 34 |  |
| Other ethnicity | 89 (<1) | 2 | 28 (36) | 36 |  |
| Changed role due to C19 |  |  |  |  |  |
| Yes | 1,510 (12) | 12 | 415 (30) | 33 | <0.001 |
| No | 11,284 (88) | 88 | 2,576 (24) | 27 |  |
| In contact with C19 patients |  |  |  |  |  |
| Yes | 6,174 (48) | 57 | 1,744 (30) | 34 | <0.001 |
| No | 6,597 (52) | 43 | 1,245 (20) | 22 |  |
| Adequate PPE |  |  |  |  |  |
| Yes | 10,018 (91) | 89 | 2,249 (24) | 27 | <0.001 |
| No | 989 (9) | 11 | 415 (46) | 49 |  |
| Feel you have adequate support from: |  |  |  |  |  |
| Line managers |  |  |  |  |  |
| Yes | 10,492 (82) | 80 | 1,853 (19) | 21 | <0.001 |
| No | 2,283 (18) | 20 | 1,135 (54) | 56 |  |
| Colleagues |  |  |  |  |  |
| Yes | 11,708 (92) | 91 | 2,468 (23) | 25 | <0.001 |
| No | 1,082 (8) | 9 | 522 (53) | 56 |  |
| Friends/family |  |  |  |  |  |
| Yes | 12,137 (95) | 95 | 2,740 (24) | 27 | <0.001 |
| No | 623 (5) | 5 | 244 (42) | 45 |  |
| Colleague died from C19 |  |  |  |  |  |
| Yes | 788 (6) | 10 | 280 (38) | 40 | <0.001 |
| No | 11,777 (94) | 90 | 2,718 (24) | 26 |  |

<sup>a</sup> Percentages may not add up to 100 due to missing data.
<sup>b</sup> Testing for differences between proportions of those meeting and not meeting cut-off
within each category.

**Table 2.**
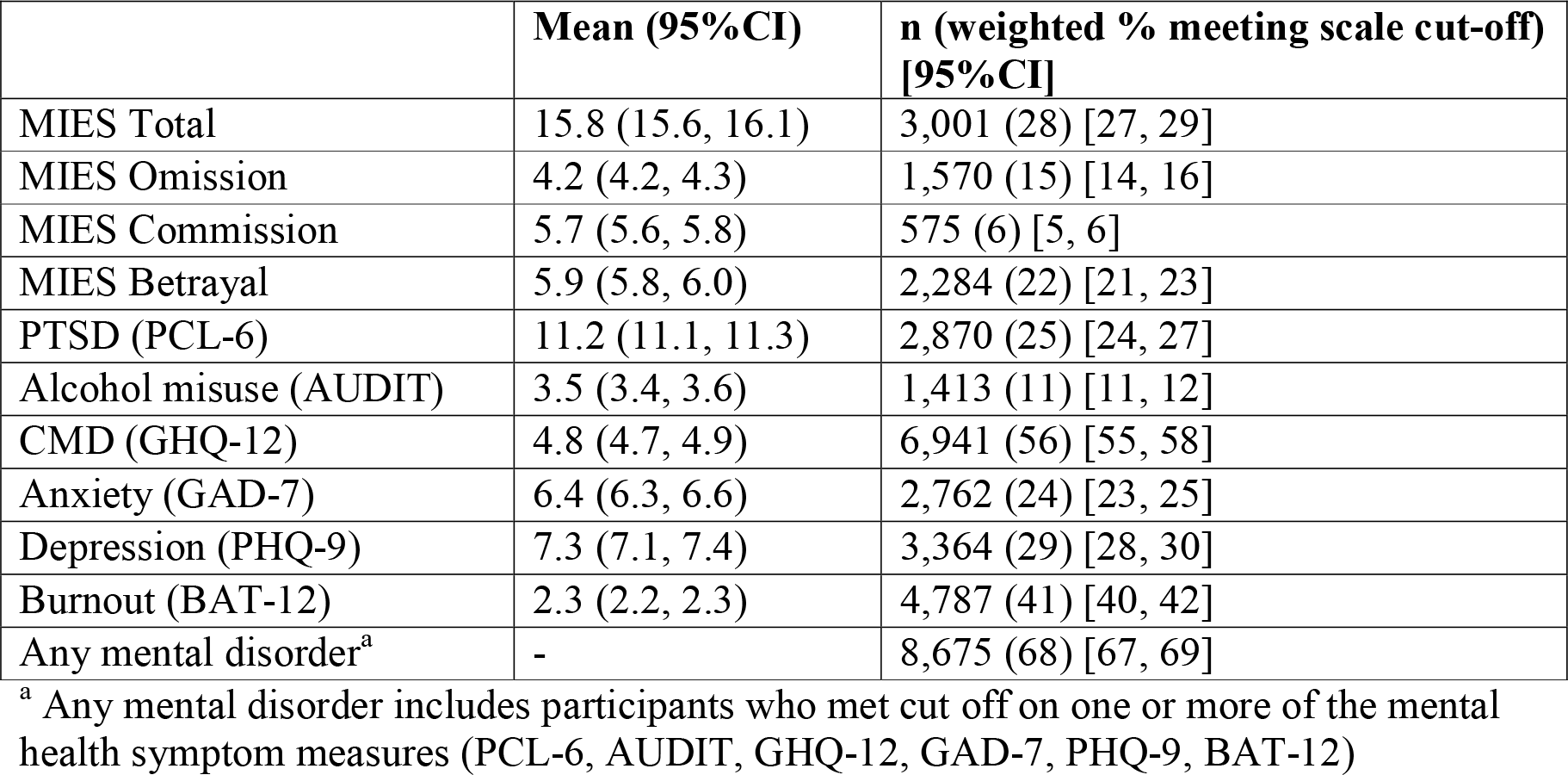
Mean scores and proportions meeting cut-off scores of measures

|  | **Mean (95%CI)** | **n (weighted % meeting scale cut-off) [95%CI]** |
| --- | --- | --- |
| MIES Total | 15.8 (15.6, 16.1) | 3,001 (28) [27, 29] |
| MIES Omission | 4.2 (4.2, 4.3) | 1,570 (15) [14, 16] |
| MIES Commission | 5.7 (5.6, 5.8) | 575 (6) [5, 6] |
| MIES Betrayal | 5.9 (5.8, 6.0) | 2,284 (22) [21, 23] |
| PTSD (PCL-6) | 11.2 (11.1, 11.3) | 2,870 (25) [24, 27] |
| Alcohol misuse (AUDIT) | 3.5 (3.4, 3.6) | 1,413 (11) [11, 12] |
| CMD (GHQ-12) | 4.8 (4.7, 4.9) | 6,941 (56) [55, 58] |
| Anxiety (GAD-7) | 6.4 (6.3, 6.6) | 2,762 (24) [23, 25] |
| Depression (PHQ-9) | 7.3 (7.1, 7.4) | 3,364 (29) [28, 30] |
| Burnout (BAT-12) | 2.3 (2.2, 2.3) | 4,787 (41) [40, 42] |
| Any mental disorder<sup>a</sup> | - | 8,675 (68) [67, 69] |

<sup>a</sup> Any mental disorder includes participants who met cut off on one or more of the mental health symptom measures (PCL-6, AUDIT, GHQ-12, GAD-7, PHQ-9, BAT-12)

As seen in Table 1, several socio-demographic characteristics were found to be associated with expressions of moral injury. Male participants (p<0.001), being a doctor or nurse (p<0.001), or those who report Asian, mixed or other ethnicity (p=0.02) were significantly more likely to meet cut off on the MIES. Expressions of moral injury were also more likely to be observed in those participants who were redeployed and changed roles during COVID-19 (p<0.001), those who reported contact with COVID-19 patients (p<0.001), and those who reported having a lack of PPE (p<0.001) were significantly more likely to meet cut off on the MIES. Experiencing a lack of support from managers, colleagues, and family members (p<0.001) was associated with meeting cut off on the MIES (p<0.001). Finally, having a colleague die of COVID-19 was significantly associated with meeting cut off on the MIES (p<0.001).

A substantial proportion of the sample met case criteria for probable mental disorders. 25% met criteria for probable PTSD, 56% for CMD, 41% for burnout and 11% met criteria for probable alcohol misuse (Table 2). The sample had a mean total MIES score of 15.8 (95%CI 15.6, 16.1), with 28% meeting MIES cut off.

Those who met cut off on the MIES were statistically significantly more likely to meet case criteria for probable PTSD, CMD, anxiety, depression, and burnout. Notably, those reported having experienced PMIES were not significantly more likely to report alcohol misuse, although this varied by event type, as reporting acts of omission (AOR 1.4, 95% CI 1.0, 1.7) and commission (AOR 1.9, 95% CI 1.4, 2.6) were significantly associated with greater alcohol misuse. Experiencing acts of omission, commission and betrayal were most strongly associated with probable PTSD (see Table 3).

**Table 3.** Odds Ratio (OR) and Adjusted Odds Ratio (AOR) for MIES total and subscales, for individuals meeting cut-off for each mental health measure

|  |  | PTSD | Alcohol<br>misuse | CMD | Anxiety | Depression | Burnout | Any MH<br>measure |
| --- | --- | --- | --- | --- | --- | --- | --- | --- |
| *MIES total*<br>*cut-off* | OR (95% CI) | 3.1 (2.8, 3.5)<br>p<0.001 | 1.2 (1.0, 1.4)<br>p=0.02 | 2.5 (2.1, 2.9)<br>p<0.001 | 2.8 (2.5, 3.1)<br>p<0.001 | 2.6 (2.4, 2.9)<br>p<0.001 | 3.1 (2.9, 3.4)<br>p<0.001 | 3.0 (2.5, 3.6)<br>p<0.001 |
|  | AOR (95%<br>CI) | **3.0 (2.4, 3.6)**<br>**p<0.001** | 1.2 (0.9, 1.5)<br>p=0.11 | **1.9 (1.6, 2.3)**<br>**p<0.001** | **2.2 (2.0, 2.5)**<br>**p<0.001** | **2.1 (1.8, 2.3)**<br>**p<0.001** | **2.3 (2.0, 2.7)**<br>**p<0.001** | **2.3 (1.9, 3.0)**<br>**p<0.001** |
| *MIES*<br>*omission* | OR (95% CI) | 2.4 (2.1, 2.9)<br>p<0.001 | 1.3 (1.1, 1.5)<br>p=0.005 | 2.0 (1.5, 2.6)<br>p<0.001 | 2.2 (1.9, 2.5)<br>p<0.001 | 2.0 (1.6, 2.5)<br>p<0.001 | 2.3 (2.0, 2.5)<br>p<0.001 | 2.6 (1.9, 3.6)<br>p<0.001 |
|  | AOR (95%<br>CI) | **2.2 (1.9, 2.6)**<br>**p<0.001** | **1.4 (1.0, 1.7)**<br>**p=0.007** | **1.6 (1.2, 2.1)**<br>**p=0.003** | **1.8 (1.4, 2.1)**<br>**p<0.001** | **1.6 (1.3, 2.1)**<br>**p<0.001** | **1.7 (1.6, 1.9)**<br>**p<0.001** | **2.2 (1.5, 3.1)**<br>**p<0.001** |
| *MIES*<br>*commission* | OR (95% CI) | 3.4 (2.7, 4.2)<br>p<0.001 | 1.7 (1.4, 2.2)<br>p<0.001 | 2.4 (1.8, 3.2)<br>p<0.001 | 3.1 (2.5, 3.8)<br>p<0.001 | 2.9 (2.4, 3.5)<br>p<0.001 | 3.0 (2.4, 3.8)<br>p<0.001 | 3.2 (2.0, 5.2)<br>p<0.001 |
|  | AOR (95%<br>CI) | **2.9 (2.2, 3.8)**<br>**p<0.001** | **1.9 (1.4, 2.6)**<br>**p=0.001** | **1.9 (1.3, 2.7)**<br>**p=0.002** | **2.5 (1.9, 3.3)**<br>**p<0.001** | **2.5 (2.0, 3.0)**<br>**p<0.001** | **2.4 (1.9, 3.0)**<br>**p<0.001** | **2.7 (1.6, 4.7)**<br>**p=0.001** |
| *MIES*<br>*betrayal* | OR (95% CI) | 3.1 (2.7, 3.5)<br>p<0.001 | 1.3 (1.0, 1.5)<br>p=0.02 | 2.8 (2.4, 3.2)<br>p<0.001 | 3.1 (2.7, 3.4)<br>p<0.001 | 2.8 (2.4, 3.3)<br>p<0.001 | 3.5 (3.1, 4.0)<br>p<0.001 | 3.3 (2.8, 4.0)<br>p<0.001 |
|  | AOR (95%<br>CI) | **2.8 (2.3, 3.3)**<br>**p<0.001** | 1.2 (0.9, 1.6)<br>p=0.13 | **2.0 (1.8, 2.3)**<br>**p<0.001** | **2.4 (2.1, 2.7)**<br>**p<0.001** | **2.2 (1.8, 2.6)**<br>**p<0.001** | **2.6 (2.2, 3.2)**<br>**p<0.001** | **2.4 (2.0, 3.0)**<br>**p<0.001** |

AOR: adjusted for age, sex, ethnicity, role, change in role, contact with COVID-19 patients, adequate PPE, adequate support from colleagues, managers, and family/friends, colleague died from COVID-19. **Bold** indicates statistically significant AOR. Any mental health measure = met criteria for one or more mental disorders.

**Table 4.**
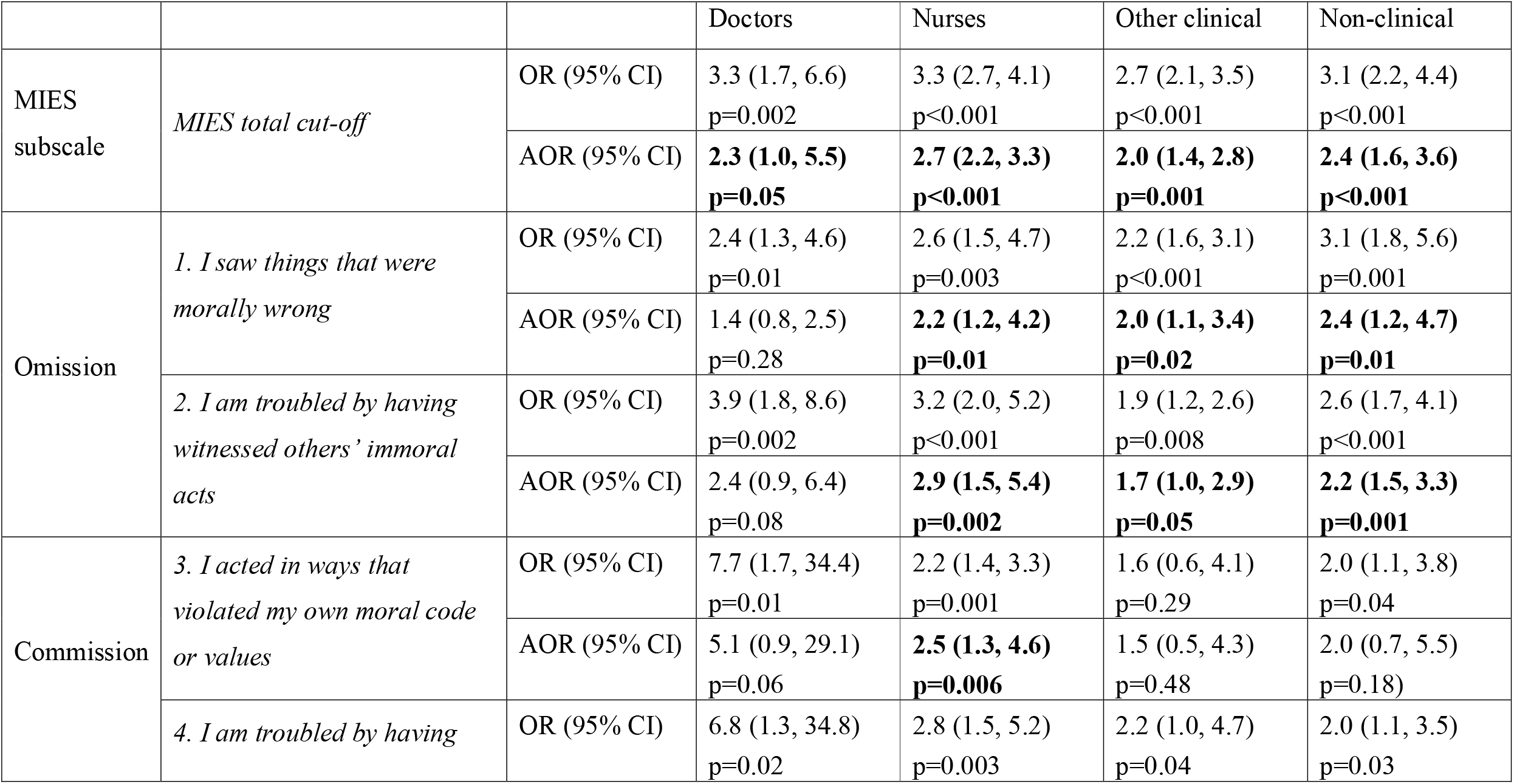
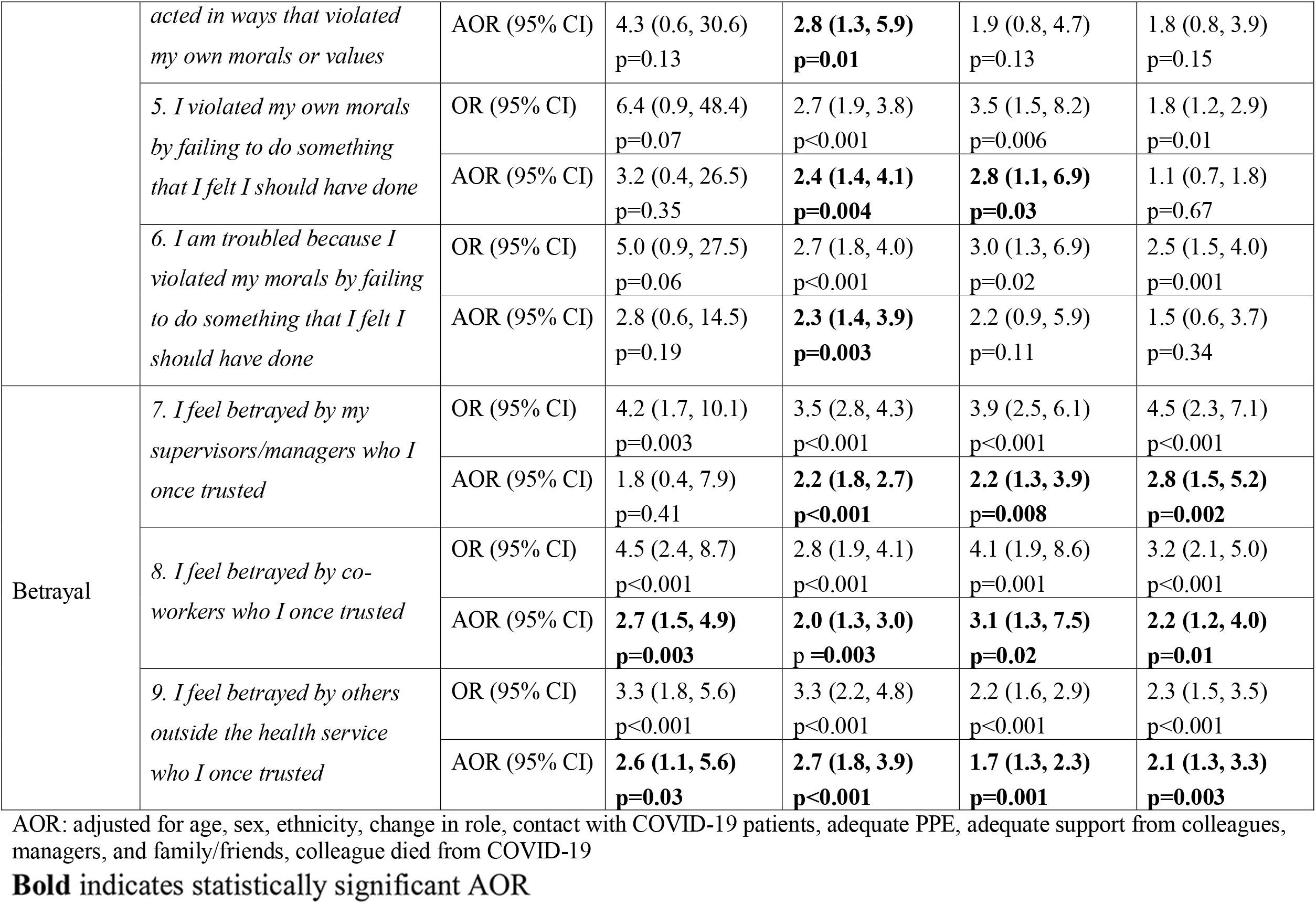
Odds Ratio (OR) and Adjusted Odds Ratio (AOR) for meeting cut-off on MIES total and subscales, for individuals meeting cut-off on one or more mental health measure, by role

When the MIES items are examined individually, different staff groups reported PMIEs differently. Doctors, nurses, other clinical staff and non-clinical staff who meet cut off for any probable mental disorder were at least twice as likely to report PMIEs compared to those who did not meet cut off on a mental health measure (see Table 4). Doctors with a probable mental disorder were more than twice as likely to report experiencing a betrayal by co-workers (item 8; AOR 2.7, 95% CI 1.5, 4.9) and betrayal by those outside the health service (item 9, AOR 2.6, 95%CI 1.1, 5.6) compared to those who did not meet case criteria for a probable mental disorder.

Nurses who met cut off for probable mental disorders were significantly more likely to report experiencing acts of omission, commission and betrayal compared to nurses who did not meet case criteria. In particular, nurses who met cut off were 2.7 times more likely to report experiencing a betrayal by others outside the health service (item 9, AOR 2.7, 95% CI 1.8, 3.9) and acting in a way that violates their morals or values (item 4, AOR 2.8, 95% CI 1.3, 5.9).

Other clinical and non-clinical staff who met cut off criteria for at least one probable mental disorder, were also significantly more likely to endorse experiences of acts of omission and betrayal as compared to those who did not meet case criteria (see Table 4).

## Discussion

This study aimed to examine the experience and impact of PMIEs in a nationally representative sample of NHS healthcare staff during the COVID-19 pandemic. Four key findings were observed. First, nearly a third of healthcare staff reported experiences of PMIEs, with acts of betrayal most frequently reported. Second, PMIE exposure was significantly associated with adverse mental health symptoms across clinical and non-clinical healthcare staff. Third, specific work-factors were significantly associated with expressions of moral injury, including being redeployed, a lack of PPE, and experiencing a lack of support. Finally, in all staff groups, those reporting symptoms of mental disorders were significantly more likely to experience PMIEs, with those in certain roles more likely to endorse specific PMIEs. Nurses reporting symptoms of mental disorders were significantly more likely to report all types of PMIEs than those without symptoms. Whereas doctors who reported mental disorder symptoms were only more likely to report betrayal events (not omission or commission). In addition to reporting betrayal events, other clinical and non-clinical staff reporting symptoms of mental health disorders were more likely than those without symptoms to report acts of omission.

That a substantial proportion (68%; 95% CI 67, 69) of the present sample met case criteria for one or more probable mental disorders, including PTSD, CMD and alcohol misuse, is consistent with previous studies [16,18,31], highlighting the current state of wellbeing in healthcare staff providing care to patients during the COVID-19 Pandemic. This research is novel in that it extends previous findings on moral injury to the UK healthcare workforce during the time of the COVID-19 Pandemic. Previous studies have found relationships between moral injury and mental disorders in other occupational groups, particularly military samples [4,5,32]. The significant relationship between experiences of moral injury and adverse mental health in UK healthcare staff found in the present study adds to the literature by illustrating that moral injury can be experienced by staff in non-military settings, and also have negative associations with mental ill-health. However, due to the cross-sectional design of the current study, we cannot infer causality. Nonetheless, the present results highlight that moral injury is associated with poor wellbeing in both clinical and non-clinical staff – particularly following acts of omission and betrayal. This highlights that moral injury is not solely applicable to clinical staff and does occur to staff who may not directly provide patient care. As safeguarding the wellbeing of staff is a priority for the NHS, these findings suggest that all staff, including those who do not work clinically, may experience moral injury and this should be considered when making provisions for staff wellbeing support.

A number of occupational factors were found to be associated with moral injury, including a lack of support from management and colleagues, perceived lack of PPE and having a colleague die of COVID-19. These findings are broadly consistent with a recent qualitative study which observed that military veterans may be more vulnerable to moral injury if they felt had a lack of support from chain of command [3]. This highlights that healthcare staff may benefit from targeted support from those in management roles, with managers prepared to have psychologically sensitive conversations with staff, offering informal support and signposting to services where necessary [10]. A lack of adequate PPE to protect staff was a problem for the NHS at the start of the pandemic in March 2020 [33]. Frankly preparing staff for the difficult tasks they will be asked to carry out in less than ideal and perhaps unsafe circumstances may potentially be protective and normalise distress [11]. Transparent discussion and acknowledgement when staff/organisational failings have occurred, as well as when staff have been placed in impossible situations, may also be experienced as validating and helpful. Those staff who re-deploy may also be most likely to benefit from (in)formal support and targeted advice. For example, staff who re-deploy should be adequately prepared for their role or, where time is lacking, be closely mentored as they begin their role. Staff should also be checked for redeployment suitability – for instance, if they are in a vulnerable group [18]– and be provided with information about the role and the risks associated so they are able to make an informed choice about re-deployment. Ideally, having redployment be explicitly voluntary may be beneficial. Where a colleague has died, it may be beneficial for managers to ensure that this is recognised and acknowledged as a impactful event and ensure appropriate support is in place. Previous studies have found that feeling satisfied by the notification of death process, where sufficient information was given in order to make sense of what happened, is related to less long-term mental distress [34]. Nonetheless, future qualitative studies to provide an in-depth understanding of the impact of PMIEs on healthcare staff wellbeing and their perceived support needs would be worthwhile in informing this process. Equally, during adversity it is possible to experience -traumatic growth [35] and this should also be examined in future healthcare staff wellbeing research.

Those with mental health problems across the healthcare sample were significantly more likely to report betrayal events. Additional research is needed to better understand *how* staff perceive the betrayal events to have occurred and by *whom* (e.g. colleague, the public, the government) to inform targeted interventions for repair. Where a betrayal event has occurred ‘in house’, reflective practice sessions, such as Schwarz rounds [36], that allow views to be expressed and solutions to be co-produced may be effective to mitigate the development of moral injury. If a betrayal is perceived to be caused by someone outside the health service, then a governmental or community approach may be more appropriate. As this study examined moral injury using the MIES, which includes items which could be interpreted in more than one way (e.g. ‘I feel betrayed by others outside the health service who I once trusted’), responses may not always reflect moral injury as it relates to staff’s occupational role. Nonetheless, recommendations of measures to manage moral injury arising outside the NHS (e.g. public enquiries) are beyond the scope of this paper

Nurses with mental disorders were more likely to report all types of PMIEs and may be more vulnerable to a broader range of PMIEs compared to other staff. It is possible that nurses may be particularly vulnerable to moral injury as they have considerable responsibilities and demands on their time, yet they also may have less ‘control’ over the care they are asked to provide despite often spending considerable time with patients and their relatives. In the general population, those most likely to experience mental disorders related to COVID-19 are often younger, female and those with children [37,38] - criteria which many nurses fit. However, given the cross-sectional nature of the study, it is unclear whether exposure to PMIEs causes mental health problems, or whether having mental health problems makes one more likely to experience PMIEs. No prospective studies of moral injury have been conducted to date and this leaves a considerable gap in our understanding of the development and impact of moral injury over time.

This study has several strengths and limitations. Amongst the strengths was the inclusion of a large and representative, weighted NHS sample. A second strength was that moral injury was examined comprehensively, exploring not only the impact of exposure on wellbeing but also at the factors associated with exposure. However, a limitation of this study is that there is no existing UK validated measure for moral injury. We used the MIES because it has been used internationally [39,40], yet as some of the MIES items include distress related to moral injury (e.g. ‘I am troubled by…’) it is possible this may have conflated effects [41]. Secondly, the MIES was only completed by participants who took part in the longer survey [18]. Evidence from the longer survey, reported in Lamb et al. [18] found women were statistically significantly more likely to complete both surveys than men, as were participants of White ethnicity, those in a relationship, those who were nurses or in non-clinical roles, and those who had not been redeployed. As nursing staff and those who were re-deployed may be at particular risk for moral injury, this may have impacted our findings. Third, the presence of mental health difficulties was evaluated using self-report measures rather than clinical interviews, which is the gold standard for mental illness assessment. Finally, the cross-sectional nature of this study cannot be overlooked, and the direction of effects is unclear.

## Conclusions

The present study illustrates that NHS healthcare staff in both clinical and non-clinical roles report exposure to PMIEs during their work in the COVID-19 pandemic. Experiences of PMIEs were significantly associated with poor mental health symptoms, which is consistent with the broader literature in other high-risk occupational groups [4]. Importantly, we identified a number of socio-demographic and occupational variables associated with expressions of moral injury, such as experiencing redeployment, which may inform the targeted advice and support offered to staff. Finally, this study found that experiences of betrayal events were especially prevalent and nurses in particular may be exceptionally vulnerable to a broader range of PMIEs than other staff. Further research is needed to develop effective interventions for moral injury, especially in healthcare settings, to safeguard the wellbeing of staff.

## Data Availability

No additional data are available.

## Acknowledgements

We wish to acknowledge the National Institute of Health Research (NIHR) Applied Research Collaboration (ARC) National NHS and Social Care Workforce Group, with the following ARCs: East Midlands, East of England, South West Peninsula, South London, West, North West Coast, Yorkshire and Humber, and North East and North Cumbria. They enabled the set-up of the national network of participating hospital sites and aided the research team to recruit effectively during the COVID-19 pandemic.

The NHS CHECK consortium includes the following site leads: Sean Cross, Amy Dewar, Chris Dickens, Frances Farnworth, Adam Gordon, Charles Goss, Jessica Harvey, Nusrat Husain, Peter Jones, Damien Longson, Richard Morriss, Jesus Perez, Mark Pietroni, Ian Smith, Tayyeb Tahir, Peter Trigwell, Jeremy Turner, Julian Walker, Scott Weich, Ashley Wilkie.

The NHS CHECK consortium includes the following co-investigators and collaborators: Peter Aitken, Anthony David, Sarah Dorrington, Rosie Duncan, Cerisse Gunasinghe, Stephani Hatch, Daniel Leightley, Isabel McMullen, Martin Parsons, Paul Moran, Catherine Polling, Alexandra Pollitt, Danai Serfioti, Chloe Simela, Charlotte Wilson Jones.

